# Genetic Relationships of Third-Generation Cephem Resistant *Escherichia coli* in Humans, Dogs and Cattle using a Novel Clonotyping Assay

**DOI:** 10.64898/2026.09.23.26363816

**Authors:** Laurel Legenza, Erin Jonaitis, John D. Lee, Evgeni V. Sokurenko, Veronika Tchesnokova, Jessica L. Hite, Song Gao, Brooke J. Olson, Thomas R. Fritsche

**Author notes:** Corresponding author: Laurel Legenza.

## Abstract

Antimicrobial resistance is a global ‘One Health’ threat connecting antimicrobial use in human health, animal husbandry, and companion animal health with concern for persistence in environmental reservoirs. Complicating this threat is the gap that exists in our understanding of the intersection of antimicrobial resistance among humans and our companion and food production animals. This study applies a visual and statistical approach to further understand antimicrobial resistance characteristics and relatedness of *Escherichia coli* found in humans, dogs and cattle. The primary study collection included third-generation cephem resistant (3GCR) *E. coli* isolates recovered from human, dog, and cattle gastrointestinal specimens and secondarily human and dog isolates recovered from infection sources, primarily urinary tract infections. We previously reported on the transferable gene targets encoding for 3GCR in colonizing isolates and here further identified the familial-like clonal group of each resistant isolate using a novel clonotype diagnostic (septatyping). Together, this approach identified distinct genetic signatures among these 3GCR isolates, many of which are peculiar to their respective host species. We found that the overlap measured by the correlation in abundance of shared clonotypes of resistant *E. coli* among hosts revealed statistically greater similarity between dogs and humans than between cattle and humans. Yet, the reservoir of AMR genes was highest in cattle and secondarily dogs. While these findings demonstrate that clonotypes of 3GCR *E. coli* in humans, dogs and cattle display significant host specificity, transient colonization from a host enriched with resistant strains may still pose considerable risk of horizontal transfer of resistance genes between hosts living in proximity to each other.

## Introduction

The global threat of antimicrobial resistance (AMR) is urgent as multidrug resistance (MDR) increasingly threatens clinical care.[1,2] Globally, the majority (73%) of antibiotics are used in animal husbandry for growth promotion, therapeutic, and prophylactic applications; use in aquaculture and horticulture is also widespread.[3] Resistance to third generation cephalosporins (3GC) in particular is highly concerning given their wide-spread use and clinical importance in both human and veterinary medicine due to potency, stability to common beta-lactamases, efficacy, broad safety profile, and low cost.

The rapid spread of extended-spectrum beta-lactamases (ESBLs), plasmidic AmpC cephalosporinases (pAmpC) and, more recently, carbapenemases in Enterobacterales in general, and *E. coli* in particular, has created a serious global health problem identified by the World Health Organization, the Centers for Disease Control and Prevention, and other leading national and international agencies and organizations.[4,5] Widespread use of 3GC for treating mastitis in dairy cattle, urinary tract, and skin infections in domestic pets and multiple infection indications in humans has been implicated in driving the spread of 3GC AMR, although the mechanisms of resistance transfer are less clear.[6] Yet, less is known about actual movement of resistance genes, either through clonal spread of *E. coli* and related species, movement of various extrachromosomal mobile genetic elements, or a combination of both.

To better understand the scope of the reservoir of AMR *E. coli* that exists in humans, dogs, and cattle, we previously screened fecal specimens in a geographically overlapping region specifically for 3GC resistance and characterized the most common resistance gene targets.^4^ In this study, we performed a novel high-resolution clonal diagnostic (septatyping, a clonotyping method based upon 7 single nucleotide polymorphisms [SNPs] occurring within the house-keeping genes fumC and fimH) on this isolate collection to better assess the overall diversity of 3GCR *E. coli* populations recovered from host species and the degree of similarity between them. A subset of temporally related *E. coli* infection isolates from humans and dogs not reported upon previously were evaluated by the same methods. Statistical measures of distributional overlap (Bhattacharyya Coefficient) and frequency similarity (Spearman’s ρ) were used to assess the degree of relatedness of 3GCR *E. coli* clonotype populations existing among the three host species. This One Health approach to better understand cross-species sharing of bacterial clones and resistance genes may assist in the design of mitigation strategies to lessen healthcare and occupational exposure risks.

## Materials and Methods

### Ethics Approval

The University of Wisconsin-Madison Institutional Review Board reviewed the study protocol and determined that this study was exempt from informed consent requirements as the study included retrospective use of existing data (IRB: 2018-1305) and followed data protection and ethical research practices.

### Specimen Sources and Collection Characteristics

This study extends our earlier publication that presented details on the prevalence of 3GC resistance found in *E. coli* colonizing the intestinal tracts of humans, dogs and cattle within our Wisconsin service area.[7] Here we included those surveillance isolates but also included 3GCR infection isolates originating from humans and dogs within the same timeframe and with a septatype result. Only isolates originating from Wisconsin were included in this analysis.

Briefly, specimen collection from the prior study consisted of consecutive and nonduplicative residual human (HumSurv, or HS; n = 2,248), dog (CanSurv, CS; n = 538) and cattle (BovSurv, BS; n = 166) fecal specimens from midwestern states submitted to the human and veterinary clinical laboratories of the Marshfield Clinic for diagnostic testing (mostly for diarrheal disease agents including for viruses, parasites, and pathogenic bacteria) during 2014. Selective chromogenic agars were used to recover *E. coli* resistant to 3GC and carbapenems from fecal specimens per manufacturers recommendations; ESBL and carbapenemase-expressing strains were included for quality control assessment. Identifications were performed using MALDI-TOF mass spectrometry and standard antimicrobial susceptibility minimal inhibitory concentration (MIC) testing was performed using the broth microdilution technique. Where more than one isolate was recovered from the selective chromogenic agars from a single specimen, both isolates were tested for resistance genes and, if different, they were included as unique isolates.

Given published performance characteristics of the FDA-cleared selective chromogenic agars, specimens from which 3GCR *E. coli* were not detected were assumed to have *E. coli* that were susceptible to this class of antimicrobial agents given the ubiquity of this species colonizing the gastrointestinal tract of mammals.

A collection of 3GCR *E. coli* isolates recovered from human (HumInfWis, HIW; n = 113) and dog (CanInf, CI; n = 87) infection sources (primarily urinary tract and, secondarily, blood stream infections and skin and soft tissue infections) and not reported previously were also studied for comparison with clonotypes recovered from host gastrointestinal tracts (all fecal samples). All Wisconsin surveillance and infection isolates were collected within a two-year time frame.

In our earlier study, isolates displaying phenotypic resistance to advanced generation cephems were tested further using a DNA microarray panel (Check-Points, Waginen, The Netherlands) to detect genes and gene variants that confer resistance to beta-lactam antimicrobics (e.g. TEM 164R, CTX-M-1 group, CTX-M-9 group, CMY-2, DHA). Terminology used was that of the microarray manufacturer; the full list of gene targets tested is in the Check-Points manual and was adapted for this publication in Supplemental Table S1.[8] Isolates displaying phenotypic 3GCR but without an identified gene target on the micro-array panel were retained in the analysis as a drug resistant isolate (ND = 3GCR mechanism not detected). Isolates displaying wildtype gene targets were excluded.

### Rapid Clonotyping

A rapid clonotyping assay was performed on all 3GCR surveillance and infection isolates using a recently described qPCR test based on the presence or absence of 7 single nucleotide polymorphisms (7-SNP typing test) occurring within the fumC and fimH genes that collectively determine the clonotype or ‘septatype’ of the bacterial isolate. This form of clonotyping has been shown to predict traditional sequence type (ST)-based clonal groups and to improve resolution through further subdivision into subclones, thereby better assessing population diversity.[9]

### Comparison of Clonal Composition of Different *E. coli* Collections

Wisconsin clonotypes of 3GCR *E. coli* were compared by host (dog vs. cattle vs. human) and to a human multisite 3GCR *E. coli* collection of clinical isolates from the United States and Europe from 2010-2016, referred to hereafter as the ‘global collection’ (HumInfGlob, HIG; courtesy of Prof. E. Sokurenko). Through this process we were able to assess overall clonal diversity among the recovered 3GCR *E. coli* surveillance and infection isolates. Furthermore, we compared the degree of similarity of clonotype populations within the three studied hosts, and between these regional hosts and the global human-source collection.

### Statistical Tests for Determining Degree of Clonal Relatedness of Host *E. coli* Populations

Distributions of clonotypes were plotted as a function of species (human, dog, cattle) and isolate type (surveillance, infection). We compared clonotype distributions across the six resulting isolate groups using two complementary analytical approaches. For each one, bootstrap resampling at the isolate level was used to construct bias-corrected and accelerated confidence intervals around each statistic.[10,11] All statistical analyses were performed in R 4.5.1.[12,13] First, we quantified similarity between each pair of subgroup distributions using the Bhattacharyya Coefficient (BC), a measure useful for comparing multinomial distributions that range from 0 (no overlap) to 1 (identical distributions).[14]

Second, we evaluated concordance in septatype frequencies using Spearman’s ρ, a nonparametric correlation suitable for skewed and/or nonlinear but monotonic relationships that ranges from -1 (perfect negative correlation) through 0 (no relationship) to +1 perfect positive correlation). To assess the sensitivity of these results to influential data points, we conducted up to three data subset analyses for each pairwise comparison:

1. the full septatype distribution;
2. datasets reduced by eliminating markers not found with a frequency of at least 2% in one of the two species being compared;
3. where not covered by step 2, further restricted datasets that additionally excluded septatype 561 (equivalent to ST131-H30), common in humans and rare to nonexistent in other species.[9]

## RESULTS

Among 2,959 Wisconsin specimens, 3GCR *E. coli* were detected in 473 specimens using chromogenic agars and microbroth dilution testing (Table 1). The prevalence of 3GCR was highest in surveillance isolates collected from cattle (52.7%, n = 88), compared to surveillance isolates from dogs (14.5%, n = 66) and humans (5.6%, n = 119). 3GCR *E. coli* isolates without a clonotype result were excluded (n = 19) from further analysis. The final sample size with paired gene target (or 3GCR phenotypic resistance but without an identified gene target) and a clonotype result was 454 isolates, including 223 human (110 surveillance, 113 infection), 143 dog (64 surveillance,79 infection) and 88 bovine (surveillance only). Among phenotypically resistant isolates with a clonotype, the microarray analysis failed to detect a mechanism of resistance in 67 isolates (14.8% of all resistant isolates;49 human, 13 dog,5 bovine). These non-detected results were retained in the analysis as mechanism not detected (ND) with their septatype result. Frequencies of septatypes and resistance mechanisms are illustrated in Figure 1.

**Figure 1.**
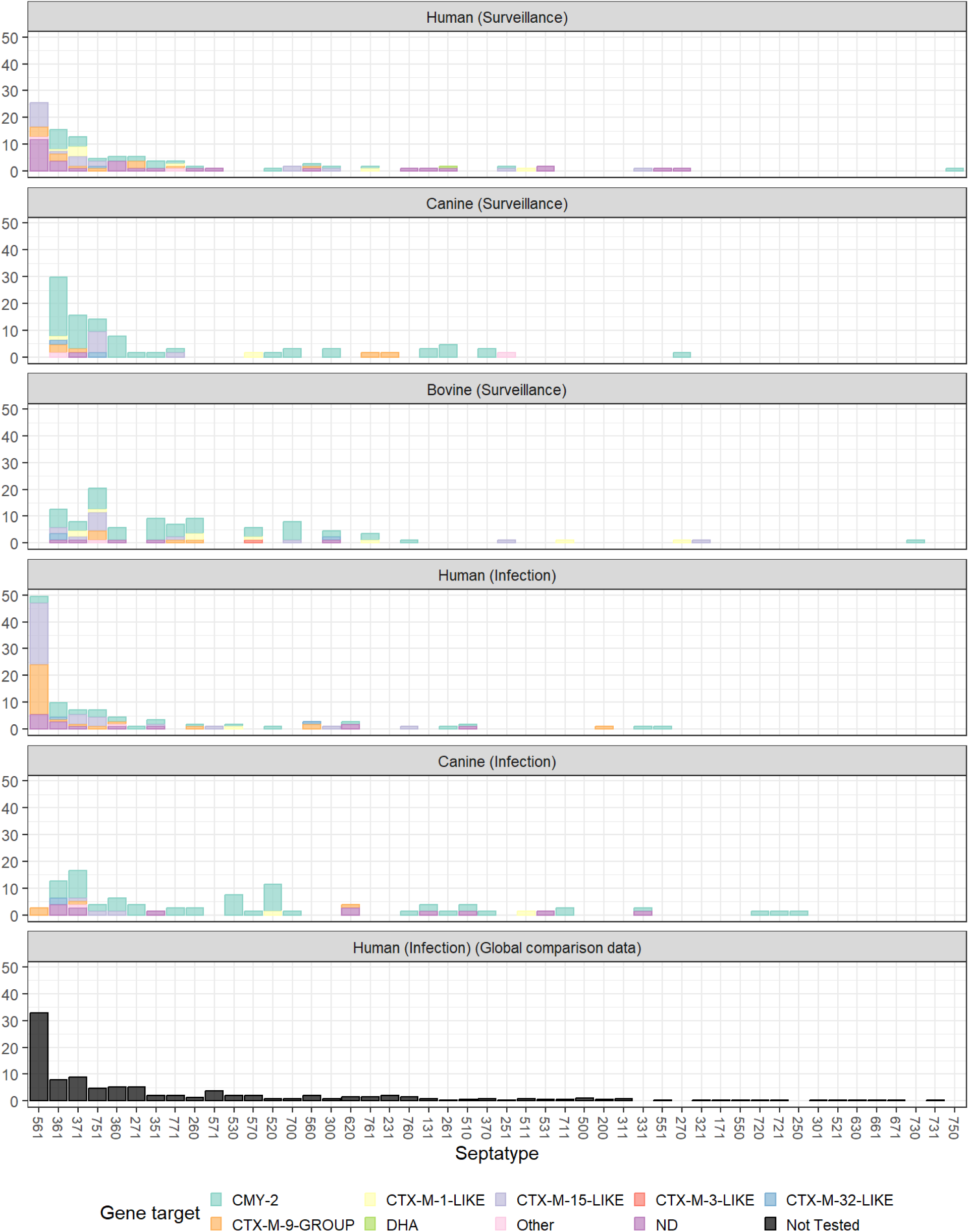
*Escherichia coli* clonal septatypes and antibiotic resistance gene target mechanisms vary by host species (human, canine, bovine) and isolate type (infection, surveillance). **Figure 1 caption:** ND: gene target not detected by assay. The colored proportions indicate the proportions of each gene target detected within each septatype by host species. The global human infection data was not tested for gene targets present.

**Table 1.** Number of surveillance and infection isolates by host species and third-generation cephem (3GCR) resistance.

| **Species** | **Isolate Type** | **3GCR Resistant** | **Susceptible** | **% 3GCR Resistant** | **Total** |
| --- | --- | --- | --- | --- | --- |
| Human | Surveillance | 119 | 2019 | 5.6% | 2138 |
| Canine | Surveillance | 66 | 388 | 14.5% | 454 |
| Bovine | Surveillance | 88 | 79 | 52.7% | 167 |
| Human | Infection | 113 | 0 | 100.0% | 113 |
| Canine | Infection | 87 | 0 | 100.0% | 87 |
| **Total** |  | **473** | **2486** |  | **2959** |

Human surveillance isolates

### Primary analyses

With all septatypes included, HumSurv isolates were most like HumInfGlob (BC_HS,HIG_ = 0.88 [0.88, 0.90]) and HumInfWis isolates (BC_HS,HIW_ = 0.85 [0.84, 0.89]). HumSurv were also similar to both CanSurv (BC_HS,CS_ = 0.74 [0.72, 0.80]) and CanInf isolates (BC_HS,CI_ = 0.72 [0.69, 0.82]). HumSurv were least like BovSurv isolates (BC_HS,BS_ = 0.67 [0.63, 0.77]). Comparing point estimates to confidence intervals, HumSurv-CanSurv and HumSurv-CanInf correlation estimates fall below human-human lower bounds, and the HumSurv-BovSurv correlation is lowest. Results using the Spearman rank correlation on the full distribution of all septatypes were largely similar, with modest, positive relationships between HumSurv and other isolates, and point estimates for the HumSurv-BovSurv correlation below correlations with CanSurv or HumInf. However, in this analysis, the HumSurv-CanInf correlation was lowest (ρ_HS,CI_ = 0.42 [0.34, 0.58]).

### Sensitivity analyses

Excluding rare septatypes, HumSurv-HumInfGlob similarity was high (ρ_HS,HIG_ = 0.97 [0.96, 1.00]), but HumSurv-CanSurv, HumSurv-CanInf, and HumSurv-BovSurv similarities dropped near to zero. However, with septatype 561 additionally excluded, strong HumSurv-CanSurv similarity emerged (ρ_HS,CS_ = 0.81 [0.60, 0.99]), but the HumSurv-BovSurv similarity remained close to zero and below HumSurv-CanSurv (ρ_HS,BS_ = 0.30 [-0.22, 0.76]), again suggesting significantly greater similarity between human and dog surveillance samples.

### Human infection-Wisconsin isolates

#### Primary analyses

HumInfWis results were similar to HumSurv. Human-human relationships were strongest (BC_HIW,HS_ = 0.85 [0.84, 0.89]; BC_HIW,HIG_ = 0.88 [0.88, 0.89]), followed by CanInf (BC_HIW,C I_= 0.66 [0.61, 0.75]), with HumInfWis-CanSurv and HumInfWis-BovSurv relationships weakest (BC_HIW,CS_ = 0.52 [0.46, 0.64]; BC_HIW,BS_ = 0.48, CI 0.43 to 0.58). Spearman full-sample results were largely similar (ρ_HIW,HS_ = 0.58 [0.52, 0.74]; ρ_HIW,HIG_ = 0.60 [0.53, 0.74]; ρ_HIW,CI_ = 0.57 [0.51, 0.75]; ρ_HIW,CS_ = 0.25 [0.092, 0.40]; ρ_HIW,BS_ = 0.26 [0.11, 0.37]).

#### Sensitivity analyses

Excluding rare septatypes, human-human correlations were higher (ρ_HIW,HS_= 0.95 [0.93, 1.00]; ρ_HIW,HIG_ = 0.92 [0.83, 0.99]) whereas human-nonhuman relationships remained modest (ρ_HIW,CI_ = 0.44 [0.055, 0.89]; ρ_HIW,CS_ = 0.48 [0.44, 0.55]; ρ_HIW,BS_ = 0.36 [0.20, 0.50]). With 561 further excluded, the HumInfWis-CanSurv and HumInfGlob-CanSurv correlations increased dramatically, and were significantly higher than HumInfWis-BovSurv.

### Canine and Bovine surveillance isolates

#### Primary analyses

CanSurv-BovSurv correlations (BC_CS,BS_ = 0.79 [0.76, 0.86]) were significantly higher than CanSurv-HumSurv (BC_CS,HS_ = 0.74 [0.72, 0.80]), CanSurv-HumInfWis (BC_CS,HIW_ = 0.52 [0.46, 0.62]), CanSurv-HumInfGlob (BC_CS,HIG_ = 0.63 [0.62, 0.67]), or CanSurv-CanInf (BC_CS,CI_ = 0.72 [0.69, 0.78]). Full-sample Spearman correlations were similar for canine-bovine and canine-human comparisons (ρ_CS,BS_ = 0.59 [0.47, 0.74]; ρ_CS,HS_ = 0.60 [0.54, 0.75]), but both were higher than correlations with infection isolates (ρ_CS,HIW_ = 0.25 [0.092, 0.40]; ρ_CS,HIG_ = 0.46 [0.41, 0.60]; ρ_CS,CI_ = 0.41 [0.30, 0.59]). Full-sample BovSurv results were similar, with BovSurv-CanSurv relationships generally stronger than between BovSurv-HumSurv, and both stronger than relationships with infection isolates (see Table 2, Figure 2).

**Figure 2.**
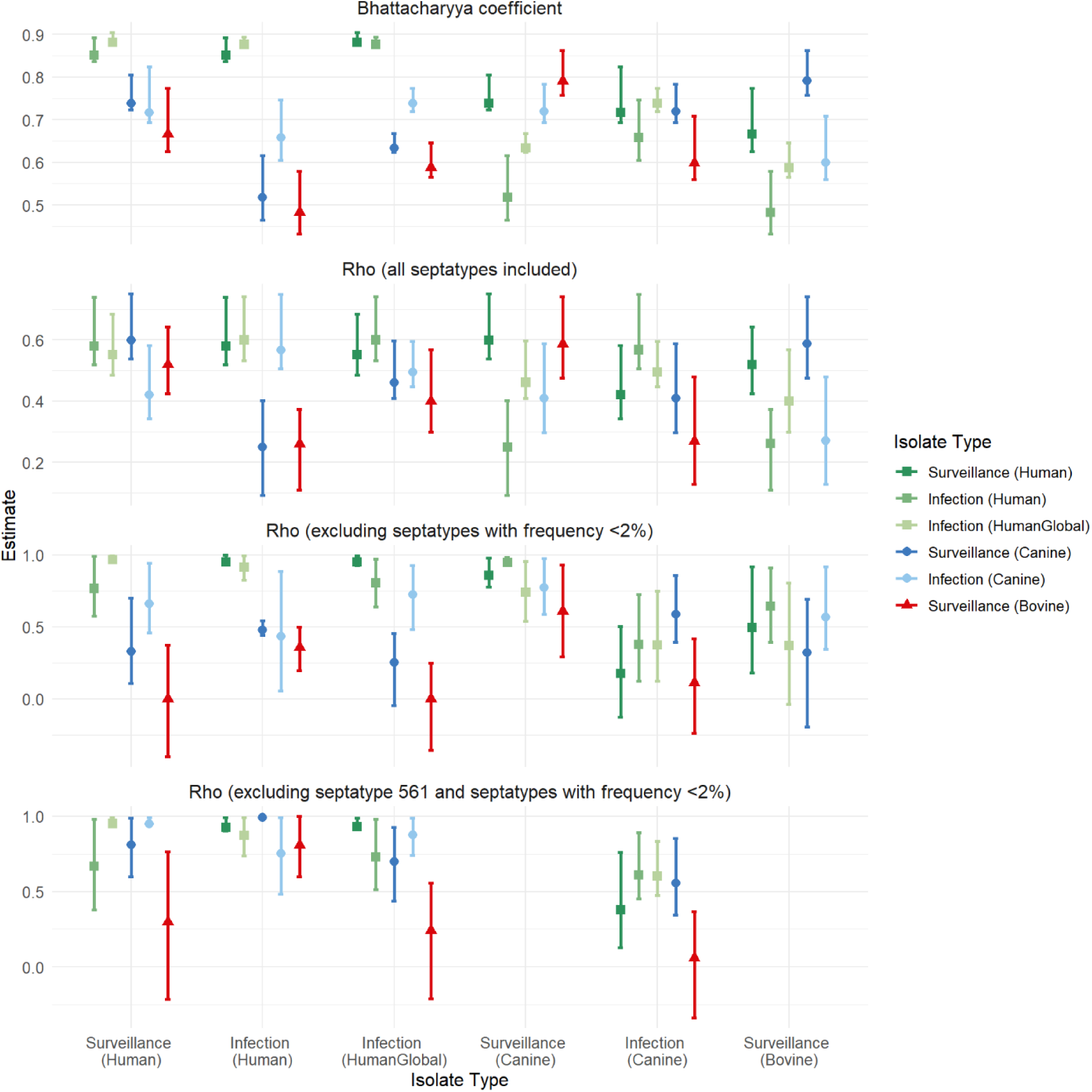
*Escherichia coli* septatype distribution concordance varies among host species (human, canine, bovine) and sample sources (infection, surveillance). **Figure 2 caption:** Concordance of septatype distributions between sample groups. X-axis groupings indicate isolate type used to limit sample inclusion, for sensitivity analyses. Error bars reflect point estimates of concordance and bootstrapped 95% confidence intervals for each comparison isolate type. Shape and hue correspond to host species (human, canine, bovine), and lightness corresponds to source (infectious or surveillance). Bovine and canine surveillance sample groups are omitted from row 4 due to redundancy with row 3, as septatype 561 had frequency <2% in these groups.

**Table 2.** Results from similarity analyses: the Bhattacharyya Coefficient and Spearman’s rank correlation comparing septatype frequency in one subgroup (species and surveillance or infection isolate type) as a function of its frequency in the other.

|  |  | Surveillance |  |  | Infection |  |  |
| --- | --- | --- | --- | --- | --- | --- | --- |
|  |  | Human | Canine | Bovine | Human | Canine | Human Global |
| Surveillance - Human |  |  |  |  |  |  |  |
| Bhattacharyya Coefficient | All septatypes included | -- | 0.74 (0.72, 0.80) | 0.67 (0.63, 0.77) | 0.85 (0.84, 0.89) | 0.72 (0.69, 0.82) | 0.88 (0.88, 0.90) |
| $\rho$ | All septatypes included | -- | 0.60 (0.54, 0.75) | 0.52 (0.43, 0.64) | 0.58 (0.52, 0.74) | 0.42 (0.34, 0.58) | 0.55 (0.49, 0.68) |
| $\rho$ | Excluding septatypes with frequency < 2% in Row Group | -- | 0.33 (0.11, 0.70) | 0 (-0.40, 0.38) | 0.77 (0.58, 0.99) | 0.66 (0.46, 0.95) | 0.97 (0.96, 1.00) |
| $\rho$ | Excluding septatype 561 and septatypes with frequency < 2% in Row Group | -- | 0.81 (0.60, 0.99) | 0.30 (-0.22, 0.76) | 0.67 (0.38, 0.98) | 0.95 (0.95, 0.99) | 0.95 (0.94, 0.99) |
| Surveillance - Canine |  |  |  |  |  |  |  |
| Bhattacharyya Coefficient | All septatypes included | 0.74 (0.72, 0.80) | -- | 0.79 (0.76, 0.86) | 0.52 (0.46, 0.62) | 0.72 (0.69, 0.78) | 0.63 (0.62, 0.67) |
| $\rho$ | All septatypes included | 0.60 (0.54, 0.75) | -- | 0.59 (0.47, 0.74) | 0.25 (0.092, 0.40) | 0.41 (0.30, 0.59) | 0.46 (0.41, 0.60) |
| $\rho$ | Excluding septatypes with frequency < 2% in Row Group | 0.86 (0.78, 0.98) | -- | 0.61 (0.29, 0.93) | 0.95 (0.95, 0.98) | 0.77 (0.59, 0.98) | 0.74 (0.54, 0.95) |
| Surveillance - Bovine |  |  |  |  |  |  |  |
| Bhattacharyya Coefficient | All septatypes included | 0.67 (0.63, 0.77) | 0.79 (0.76, 0.86) | -- | 0.48 (0.43, 0.58) | 0.60 (0.56, 0.71) | 0.59 (0.57, 0.65) |
| $\rho$ | All septatypes included | 0.52 (0.43, 0.64) | 0.59 (0.47, 0.74) | -- | 0.26 (0.11, 0.37) | 0.27 (0.13, 0.48) | 0.40 (0.30, 0.57) |
| $\rho$ | Excluding septatypes with frequency < 2% in Row Group | 0.50 (0.18, 0.92) | 0.32 (-0.19, 0.69) | -- | 0.65 (0.40, 0.91) | 0.57 (0.35, 0.92) | 0.37 (-0.037, 0.81) |
| Infection - Human |  |  |  |  |  |  |  |
| Bhattacharyya Coefficient | All septatypes included | 0.85 (0.84, 0.89) | 0.52 (0.46, 0.62) | 0.48 (0.43, 0.58) | -- | 0.66 (0.61, 0.75) | 0.88 (0.88, 0.89) |
| $\rho$ | All septatypes included | 0.58 (0.52, 0.74) | 0.25 (0.092, 0.40) | 0.26 (0.11, 0.37) | -- | 0.57 (0.51, 0.75) | 0.60 (0.53, 0.74) |
|  |  | Human | Canine | Bovine | Human | Canine | Human Global |
| $\rho$ | Excluding septatypes with frequency < 2% in Row Group | 0.95 (0.93, 1.00) | 0.48 (0.44, 0.55) | 0.36 (0.20, 0.50) | -- | 0.44 (0.055, 0.89) | 0.92 (0.83, 0.99) |
| $\rho$ | Excluding septatype 561 and septatypes with frequency < 2% in Row Group | 0.93 (0.90, 0.99) | 0.99 (1.00, 1.00) | 0.81 (0.60, 1.00) | -- | 0.75 (0.48, 0.99) | 0.87 (0.74, 0.99) |
| Infection - Canine |  |  |  |  |  |  |  |
| Bhattacharyya Coefficient | All septatypes included | 0.72 (0.69, 0.82) | 0.72 (0.69, 0.78) | 0.60 (0.56, 0.71) | 0.66 (0.61, 0.75) | -- | 0.74 (0.72, 0.77) |
| $\rho$ | All septatypes included | 0.42 (0.34, 0.58) | 0.41 (0.30, 0.59) | 0.27 (0.13, 0.48) | 0.57 (0.51, 0.75) | -- | 0.50 (0.45, 0.60) |
| $\rho$ | Excluding septatypes with frequency < 2% in Row Group | 0.18 (-0.13, 0.51) | 0.59 (0.40, 0.86) | 0.11 (-0.24, 0.42) | 0.38 (0.12, 0.73) | -- | 0.37 (0.13, 0.75) |
| $\rho$ | Excluding septatype 561 and septatypes with frequency < 2% in Row Group | 0.38 (0.13, 0.76) | 0.56 (0.34, 0.85) | 0.061 (-0.34, 0.37) | 0.61 (0.45, 0.89) | -- | 0.60 (0.48, 0.83) |
| Infection – Human Global |  |  |  |  |  |  |  |
| Bhattacharyya Coefficient | All septatypes included | 0.88 (0.88, 0.90) | 0.63 (0.62, 0.67) | 0.59 (0.57, 0.65) | 0.88 (0.88, 0.89) | 0.74 (0.72, 0.77) | -- |
| $\rho$ | All septatypes included | 0.55 (0.49, 0.68) | 0.46 (0.41, 0.60) | 0.40 (0.30, 0.57) | 0.60 (0.53, 0.74) | 0.50 (0.45, 0.60) | -- |
| $\rho$ | Excluding septatypes with frequency < 2% in Row Group | 0.95 (0.93, 0.99) | 0.25 (-0.045, 0.46) | 0 (-0.36, 0.25) | 0.80 (0.64, 0.97) | 0.72 (0.48, 0.93) | -- |
| $\rho$ | Excluding septatype 561 and septatypes with frequency < 2% in Row Group | 0.93 (0.92, 0.99) | 0.70 (0.44, 0.93) | 0.24 (-0.21, 0.56) | 0.73 (0.51, 0.98) | 0.88 (0.74, 0.99) | -- |

#### Sensitivity analyses

Results excluding rare septatypes were similar for CanSurv-BovSurv (ρ_CS,BS_ = 0.61 [0.29, 0.93]), but increased for CanSurv-HumSurv (ρ_CS,HS_ = 0.86 [0.78, 0.98]) and CanSurv-HumInfWis (ρ_CS,HIW_ = 0.95 [0.95, 0.98]). Sensitivity analyses with bovine isolates produced confidence intervals too wide to draw firm conclusions (Table 2, Figure 2).

## Discussion

Our study adds to the limited understanding of the intersection of human-animal AMR epidemiology, specifically, contributing data that answers: to what degree are colonizing and infecting resistant *E. coli* isolates similar among humans and our companion and domestic animals? To address this question, we examined similarities between 3GCR surveillance and infection isolates collected from humans, dogs and cattle (surveillance only) in Wisconsin and compared these isolates to a human global 3GCR infection collection from across the United States and Europe.

Overall, human surveillance and infection isolates were similar in both the Wisconsin-specific and global collections. Human surveillance isolates were statistically more similar to dog isolates than to cattle isolates, except for rare and human-only septatypes. Cattle surveillance isolates, however, were statistically less similar to human infection isolates than to isolates from other hosts, although in sensitivity analyses retaining only common isolates, this difference did not persist. The similarity in *E. coli* clonotypes among humans and dogs is likely related to the degree of cohabitation of humans and these pets, and adaptation to each other’s resident microbiome. The overlap of gene targets among distinct patterns of septatype frequency within each host suggests that gene targets may be moving between septatypes via plasmid exchange, while resident septatypes are more species specific.[15,16]

Our iterated similarity analyses by each host and isolate type revealed that similarity of septatypes among each host is sensitive to inclusion criteria. The overwhelming influence of the highly prevalent 561 septatype obscures an underlying similarity between human and dog septatypes, particularly in surveillance samples, suggesting non-561 *E. coli* are commonly shared between humans and canines. Point estimates for human and canine infections reflected a similar pattern. When comparing human and cattle surveillance isolates, point estimates tended to be lower than for other comparisons involving either one. This was seen both for human surveillance samples (more similar to canine than bovine surveillance) and for human infection samples (which related less strongly to bovine surveillance than canine surveillance, with rare septatypes excluded). However, the wide confidence intervals and sensitivity to inclusion criteria for the most restricted comparisons each suggest caution in interpretation.

Our data (Figure 1) highlight overlap of septatypes with the same gene targets among humans, dogs and cattle. Septatypes 361, 371, 751, and 360 contained overlapping gene targets in all three hosts, especially the clinically relevant CTX-M15-like gene target. CMY-2 also occurs within *E. coli* isolates collected from humans and animals, showing that multidrug-resistant gene targets exist in both infection and carriage isolates. However, some of the common septatypes in our study correlate to more than one distinct clonotype, and the most common human septatype, 561 (=ST131,H30), was rare in canines, and not identified at all in bovines. Thus, results support the possibility of cross-species transmission of the 3GCR gene targets between host-specific septatypes via other mechanisms (extrachromosomal elements), more so than the *E. coli* themselves becoming established in a new host.

A transient colonizing *E. coli* septatype from a human host could transfer its 3GCR gene target (e.g., CTX-M9 group) to an established *E. coli* septatype within a dog host through plasmid exchange. In our results, the few human-like 561 septatypes found in dogs carried the more common CTX-M9 group – these isolates may have been transmitted from a human reservoir. It appears less likely that host-specific septatypes are translocating to alternative hosts per the distribution of septatypes observed and lack of overlap in cattle *E. coli* compared to dog and human hosts. Still, the largest reservoir of 3GCR gene targets by proportion of *E. coli* remains in cattle.

Our findings build on existing One Health literature. Prior research demonstrates that *E. coli* from cattle are genetically distinct from *E. coli* causing human bloodstream infections, and that ESBL-producing *E. coli* in pigs are genetically distinct from those found in pig farm worker hosts.[17,18] In a study of dogs and their owners, 46.4% of dog–owner pairs had identical patterns of presence or absence of six antimicrobial resistance genes.[19] Similarity in MDR *Klebsiella* between humans and companion animals also demonstrates that potential role of companion animals in maintaining a resistance gene pool existing in parallel to that of humans.[20] Our results may reflect, in part, on drug usage patterns evident in biomedical and veterinary medicine.[21,22] The overlap of 3GCR gene targets corresponding to clinically important antimicrobials underscores the need for prudent management of antimicrobials in humans and animals.[23]

### Limitations

Results described here are somewhat limited in their generalizability to areas beyond the Upper Midwest, USA. Results with human isolates do, however, show high similarity to the global human uropathogenic 3GCR *E. coli* collection. We do not have an indicator of animal husbandry or pet ownership connections for the human isolates, although farming and pet ownership are common in Wisconsin. As the movement of resistance genes among plasmids and between *E. coli* isolates in different hosts is complex and difficult to trace, more precisely matched spatiotemporal sampling is needed.[24] Our results alone do not demonstrate transmission occurring between host species, similar to scoping review findings.[25] Still, similarities in clonotype and gene target across species suggest probable movement of organisms and transfer of gene targets. More complete genetic sequence data and controlled studies with rigorous sampling regimes are needed to explore transmission dynamics and directionality.

### Future Directions

These results are a step on the translational spectrum, ultimately towards informing mitigation strategies and empiric treatment decisions at the point of care. Further research is needed as to how septatype determination could inform antibiotic resistance predictions and guide empiric treatment given septatype data in some instances are likely more predictive of antimicrobic susceptibility than traditional antibiograms.[9] CTX-M9 isolates in humans, for example, remain susceptible to ceftazidime and cefepime, while CTX-M1 group isolates express high rates of resistance to these antibiotics across species.[7] Thus, treatments that might otherwise be excluded in the context of suspicion for 3GCR could be considered with data on the predominant clonotypes and gene targets within a geographic region or host species.

## Conclusions

Using rapid clonotype diagnostic testing along with similarity analyses, we identified strong relatedness in clonotypes between humans and dogs, and limited relatedness between humans and cattle, and between dogs and cattle. We also documented a high degree of overlap in the genes conferring 3GCR across *E. coli* colonizing humans, dogs, and cattle, though the possibility of cross-species transmission requires further exploration. Investigation of AMR transmission using whole genome sequencing of *E. coli* recovered from humans and animals would be an appropriate next step.

## CRediT Authorship Contribution Statement

**LL:** Conceptualization, Data curation, Formal analysis, Funding acquisition, Methodology, Visualization, Writing - Original Draft, Writing - Review & Editing; **EJ:** Formal analysis, Methodology, Visualization, Writing - Review & Editing; **JDL:** Formal analysis, Methodology, Visualization, Writing - Review & Editing; **EVS:** Data curation, Methodology, Writing - Review & Editing; **VT:** Data curation, Formal analysis, Methodology, Writing - Review & Editing; **JLH:** Methodology, Visualization, Writing - Review & Editing; **SG:** Methodology, Visualization, Writing - Review & Editing; **BJO:** Data curation, Methodology, Writing - Review & Editing; **TRF:** Conceptualization, Formal analysis, Funding acquisition, Methodology, Writing - Review & Editing

## Data Availability

The datasets generated and analyzed during the current study are not publicly available due to the terms of data collection and inclusion of patient data. However, de-identified aggregate data may be available from the corresponding author upon reasonable request.

## Acknowledgments

We thank Jim Lacy, Kyle McNair, and Ethan Lucas for their technical assistance.

